# A language model framework for sequence modeling of EHR audit logs to characterize clinician-EHR interactions

**DOI:** 10.64898/2026.06.24.26356449

**Authors:** Seunghwan Kim, Sunny Lou, Adam Cobb, Susmit Jha, Thomas Kannampallil

## Abstract

**Objective:** Electronic health record (EHR) audit logs capture clinician-EHR interaction patterns, but most audit log research relies on aggregated measures (e.g., total time). We investigated how audit logs could be modeled using large language model (LLM) architectures to learn underlying workflow sequences during clinician-EHR interactions.

**Materials and Methods:** Using >295 million EHR-based audit log actions from inpatient settings spanning 2019 to 2024, we fine-tuned Llama-3-8B under two encodings: (1) symbolic field-based tokens and (2) semantic natural language audit log action descriptions. A first-order Markov model, which used only the immediate prior action, served as baseline minimal context comparator. Model representation was assessed using next-action prediction accuracy in an early-period test set and two temporally distinct out-of-sample (OOS) periods.

**Results:** In the early period test set, the semantic LLM achieved the highest accuracy (0.7418, 95%CI [0.7415–0.7420]) compared to the symbolic LLM (0.3838, 95% CI: 0.3836–0.3840) and Markov baseline (0.4553, 95%CI [0.4551–0.4555]). The semantic approach was also robust to temporal drift in EHR interaction patterns seen in the two OOS periods (semantic LLM vs Markov accuracy, OOS-1: 0.6509 vs 0.3169; OOS-2: 0.6232 vs 0.2648).

**Discussion and Conclusions:** Semantic LLM relying on audit log action descriptions yielded the highest next-action prediction accuracy, and demonstrated robustness to temporal drift, suggesting that longer sequential context and semantic action descriptions may improve audit log-based sequence modeling. These findings support further development of semantic sequence models for audit log research, including task identification, automated workflow characterization, and safety-focused analyses of clinician-EHR interaction patterns.

## Introduction

Modern clinical work is primarily documented via the electronic health record (EHR).^1,2^ As such, a clinician’s work activities – ordering medications, reviewing results, writing notes – are mediated through EHR interactions. Digital traces of these interactions are recorded as metadata for regulatory purposes, and are broadly referred to as audit logs.^3^ Audit logs capture an ordered sequence of a clinician’s activities within the EHR and provide a record of clinical work;^4,5^ therefore, these audit log-based action sequences are shaped by clinicians’ professional expertise, patient situations, clinical task requirements, and EHR user interfaces.^6–9^

Prior research using audit logs has largely focused on characterizing EHR use patterns based on aggregated measures (e.g., total time spent on a task, number of clicks).^10–12^ Other studies aggregated audit logs to measure proxy constructs such as clinician workload^13–22^ (e.g., total number of actions in a day), cognitive burden^23,24^ (e.g., patient switching), or to assess the relationship between these constructs and clinician-centered outcomes such as burnout,^14,25–30^ risk for ordering errors,^23,31,32^ or reimbursement.^33^

There is limited research characterizing the sequential and structural patterns of EHR audit log action sequences, which may provide insights into a clinician’s work activities (e.g., clinical tasks such as medication ordering) or workflows. For example, techniques from natural language processing (NLP) have been applied to determine the “grammar” (i.e., a pattern of sequences similar to language) of clinical work activities.^34–40^ Most of these studies have relied on embedding short-run sequences or individual actions within a sequence, without accounting for the temporal progression or dependencies within action sequences.

Previously, we conceptualized long-range clinician-EHR interaction sequences as a language,^41,42^ allowing us to leverage large language model (LLM) architectures to ascertain trajectories of clinical workflow; this framework, which we referred to as “EHR actions-as-language,” provided a rich grammatical lens for assessing longer sequences of clinician activity patterns.^8^

In this paper, we systematically investigated the use of LLM architectures to characterize clinician-EHR interaction sequences as language-based action representations of clinical work activities. Specifically, we compared symbolic (field-based) and semantic (word-based) LLM representations for next-action prediction and evaluated the performance robustness to temporal shifts in EHR use. LLM performance was evaluated compared to a first-order Markov model – a baseline approach that only used information about the immediately prior action – to measure the impact of the LLM’s ability to learn from long-range preceding actions, which we hypothesized would encode greater context for the clinician’s work.

## Materials and Methods

### Study Setting and Data

This study was conducted at the inpatient settings of a large academic health system in St. Louis, Missouri. Data were collected between January 1, 2019, and June 1, 2024, and included EHR audit logs preceding inpatient ordering events. Specifically, we collected audit log actions 30 minutes prior to an inpatient medication or procedure order. Clinician characteristics (e.g., role, specialty, department), associated patient encounters, and orders were retrieved from Epic EHR’s (Verona, WI) Clarity database.

Our choice to use action sequences associated with ordering was based on two factors: first, ordering is a discrete clinical activity that has direct implications for patient care, with considerable evidence of associated errors.^43,44^ Second, in inpatient settings, medication ordering is one of the most common activities, with a single patient potentially having >20 orders per day.^45^

We retrieved audit log actions 30 minutes prior to each ordering event from the ACCESS_LOG table. Each audit log action included a timestamp, and ID and description of the action performed (a METRIC_ID and METRIC_NAME), a de-identified patient identifier (Patient MRN, encounter ID), and a de-identified clinician identifier.

This study was approved by the Institutional Review Board of Washington University (IRB#202209008) with a waiver of informed consent as the study was considered minimal risk.

### Data partitioning

Ordering events and associated audit log action sequences were partitioned using a temporal data-splitting strategy into 3 groups: early-period (2019/01–2020/07), and two out-of-sample (OOS) cohorts (OOS-1: 2021/06–2022/05 and OOS-2: 2023/06–2024/05) used to evaluate performance robustness to temporal drift in the EHR interface and workflows.

The early-period data set was designated for use in model development and was randomly split in a 70:10:20 ratio for training, early stopping, and evaluation (hereafter referred to as *early-period test set*).

Each OOS cohort was separated from the prior period by approximately 1-year. OOS datasets were not used during model development and were reserved exclusively for evaluation. This design allowed for the assessment of model robustness under potentially different distributions of EHR activity. A schematic of the data partitioning and evaluation strategy is provided in Figure 1.

**Figure 1.**
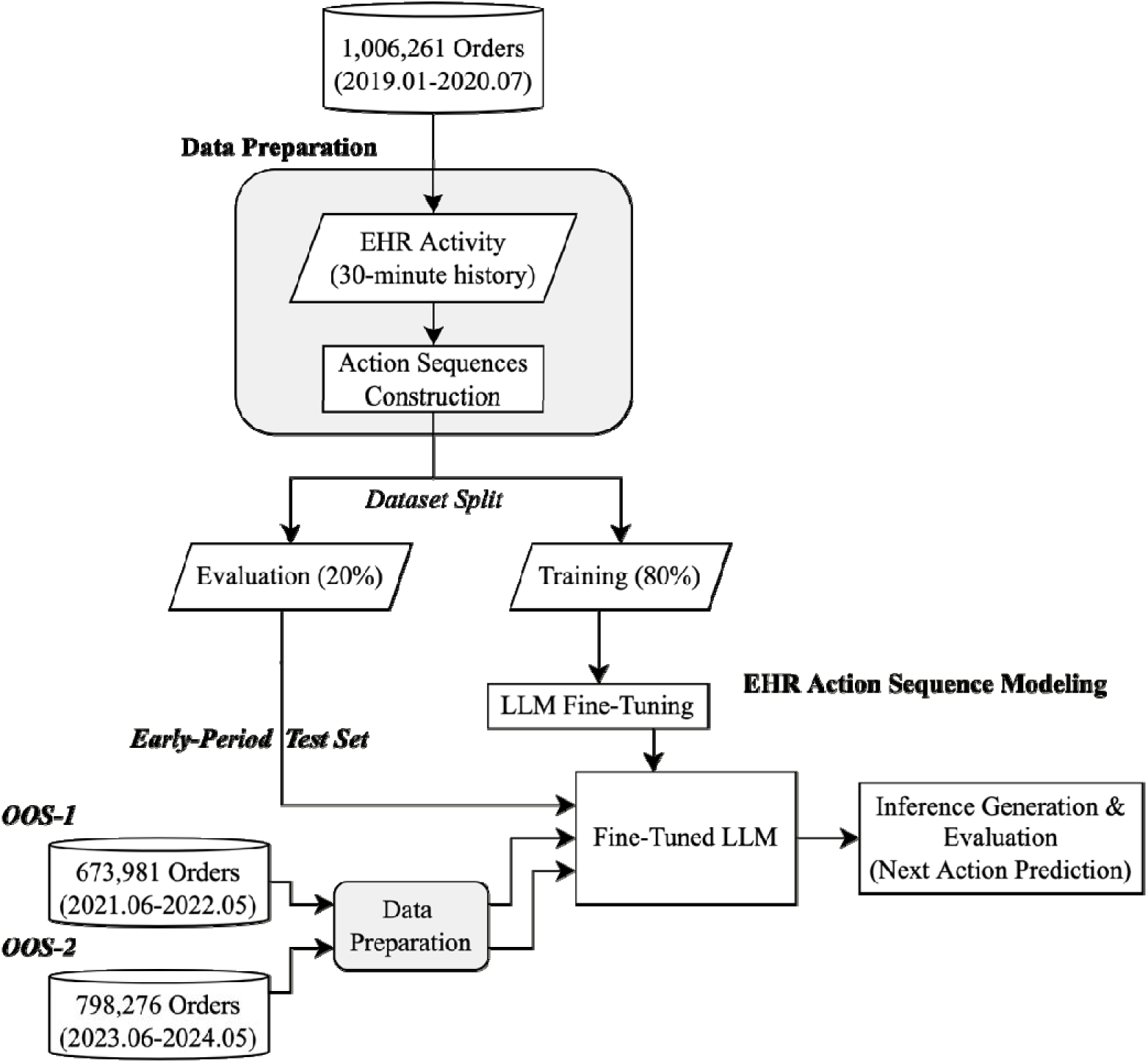
Overview of the experimental pipeline for modeling clinician interaction sequences. The EHR action history of 1,006,261 orders in the previous 30 minute were collected from 2019.01–2020.07 and structured into action sequences. The samples were then split for model fine-tuning and within-period evaluation. An LLM was fine-tuned to predict the next EHR given previous actions, at each time step within sequences. The model’s inference capabilities were initially tested on a held-out set coming from the same period of the fine-tuning set (‘early-period test set’). Additional external validation is conducted using EHR action sequences from two separate sets from later time periods (OOS-1: 2021.06–2022.05, OOS-2: 2023.06–2024.05) to test model generalizability.

The sequence of EHR audit log actions leading up to each order was used for modeling. For the LLM-based approaches, each sequence was truncated to its most recent tokens (and padded as needed) to fit a fixed context-window size (e.g., 512 tokens), chosen based on computational constraints. Because truncation retained only the most recent tokens, actions occurring earlier in long sequences were dropped before tokenization.

For the symbolic representation, each action corresponds to approximately one token, so a 512-token window spans roughly 512 actions and few action sequences exceeded this length to require truncation; for the word-based representation, each action description fragments into multiple subword tokens, so the same context window spans far fewer actions (approximately five-fold fewer), and a larger share of the earliest actions in long sequences fell outside the window and were therefore neither modeled nor evaluated. As a result, on each scored action, the word-based model had access to fewer preceding actions than the field-based model; this places the word-based representation at a context disadvantage rather than an advantage in the comparisons that follow.

### Action-as-language LLM pipeline

Figure 2 presents a schematic overview of the action-as-language modeling pipeline. The pipeline included three core components: (1) a tokenizer that converted structured action sequences into mathematical vectors, (2) transfer learning using weights from a pretrained LLM (Llama-3^46^), and (3) fine-tuning the model on the training data using parameter-efficient techniques such as model quantization and low-rank adaptation (QLoRA^47,48^). The model pipeline used EHR action sequences as input; for each action in the sequence, the model was trained to predict that action given the previous actions (Supplementary Table A1).

**Figure 2.**
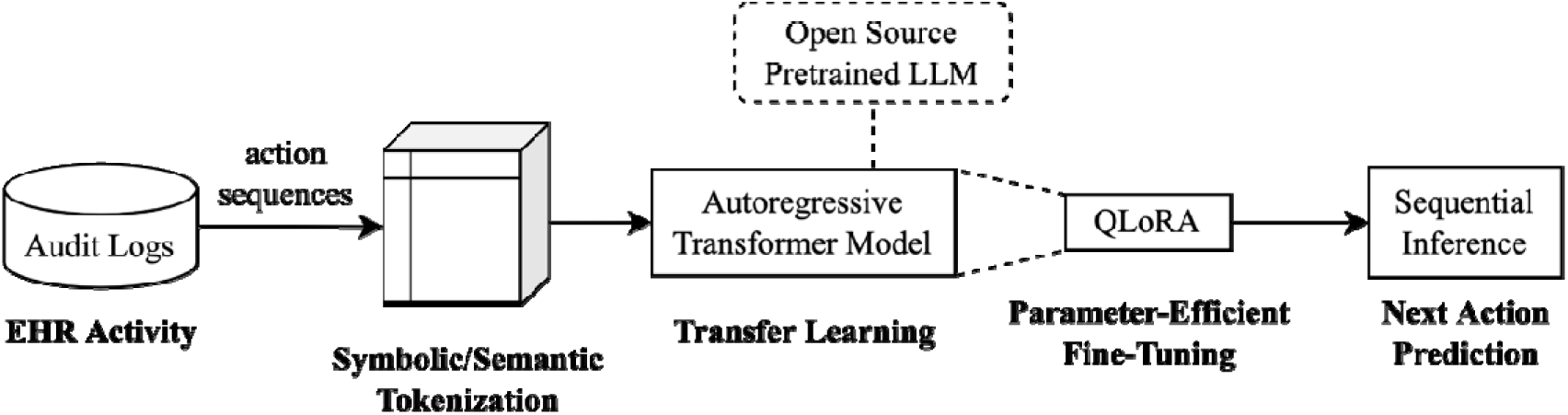
Overview of the LLM pipeline that learns and infers next EHR action in a sequence at each step. The core components are: (i) sequence tokenization of structured audit logs, (ii) transfer learning from pretrained LLMs (e.g., Llama-3), and (iii) parameter-efficient fine-tuning using model quantization and low-rank adaptation (QLoRA).

**Figure 3.**
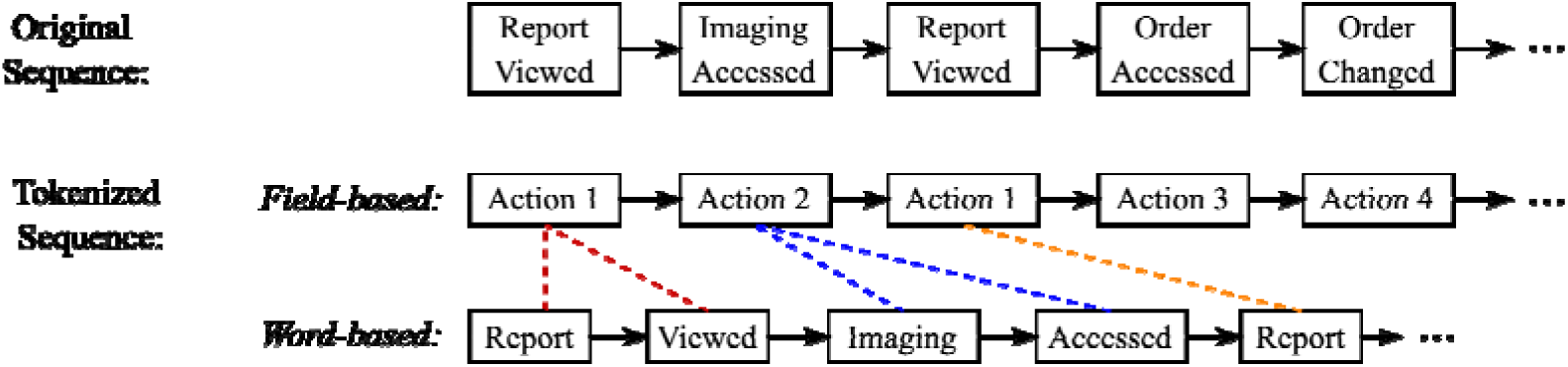
Example of field-based (symbolic) versus word-based (semantic) tokenization of clinician action sequences. The top row shows original action descriptions. The middle row shows symbolic tokenization (field-based), preserving action sequence structure. The bottom row shows word-based tokenization, enabling semantic modeling while fragmenting individual actions into multiple natural language tokens.

Within EHR audit logs, actions are recorded as discrete tokens (i.e., a METRIC_ID), where each type of EHR action that displays or modifies patient information has its own unique identifier. However, to make these actions more human-readable, EHR vendors have created descriptive labels for each METRIC_ID (METRIC_DESCRIPTION, e.g., “Report with patient data viewed”). Although these descriptive labels can provide insights into the audit log action being performed, they can be inconsistent and are often difficult to interpret without clinical and EHR expertise. Given this, we modeled EHR audit log actions in two specific ways: (a) a field-based (symbolic) representation, where each action was embedded using its unique *symbolic* token (e.g., METRIC_ID = 19330) using a custom tokenizer; this approach forced the model to focus on the structural role of each action relative to its neighbors, while abstracting the descriptive labels; and (b) a word-based (semantic) representation, where each action was represented using its full text description (e.g., “Report with Patient Data Viewed”), tokenized using a pre-specified natural language tokenizer, and embedded as English sub-words; this preserved the semantic detail of the action descriptions and allowed the LLM to leverage its pretrained language understanding. However, the resulting sub-word tokenization fragmented a single action into multiple tokens, and therefore at inference evaluation, we mapped the generated text to the closest real action description. Specifically, generated descriptions were matched against the full vocabulary of observed action descriptions using TF-IDF character n-gram embeddings (3–5 grams) and cosine similarity, with the nearest valid description selected as the predicted action. For example, an LLM’s prediction of “Report Reviewed”, which is not a valid METRIC_DESCRIPTION, was mapped to the nearest real label, “Report with Patient Data Viewed” (see Supplemental Material A2 for more detail).

In the modeling phase, we adopted a transfer learning approach by leveraging publicly available pretrained LLMs (e.g., Llama 3), which encode linguistic structure and contextual reasoning capabilities. We used the Llama-3-8B architecture^49^ and initialized the model with its pretrained weights, then fine-tuned it on EHR action sequences to predict the next EHR action at each time step in the sequence. Fine-tuning was performed separately under two representational schemes. In the word-based approach, we used the model’s default pretrained tokenizer to encode each action’s original text description directly. In the field-based approach, we extended the tokenizer with custom symbolic vocabulary.

All fine-tuning was conducted using the SFTTrainer module in Hugging Face’s Transformers Reinforcement Learning (TRL) library,^50^ with model loss and accuracy logged via Weights & Biases^51^ throughout training. Both tokenization strategies were trained under identical optimization settings to ensure a fair comparison in the next-action prediction task. Additional training details are provided in Supplementary Material A1.

Code for our modeling pipeline is available at: https://github.com/thomas-k-wustl/EHR_action-as-language.

### Model evaluation

#### Baseline comparator

The primary comparison involved the two input encoding strategies: field-based and word-based tokenization. As a non-contextual reference point, we implemented a first-order Markov baseline using bigram transition probabilities estimated from the full training set. This model predicted the next action solely from the immediately preceding action, and thus is not able to leverage any long-range action patterns. A supplementary figure (Appendix Figure A1) illustrates the contrast between the LLM’s ability to learn long-range dependencies and the Markov model’s one-step transitions. Supplemental Table A2 summarizes the model configurations evaluated in this experiment.

#### Evaluation metrics

Model performance was evaluated by measuring: (1) accuracy at the next-action prediction task, (2) accuracy at top-5 candidates (accuracy@top-5), referring to frequency with which the correct action was within the top 5 most likely actions identified by the models. We selected next-action prediction as a benchmark task as it provides an objective way to compare representations of audit-log sequences without requiring manually labeled workflow segments. Performance was compared to the baseline Markov approach on the early-period test set and the two temporal validation OOS cohorts. Detailed descriptions of the metrics are included in the Supplementary Material (Section A2).

For each metric, we computed bootstrapped 95% confidence intervals (CIs) using 1,000 resampled iterations at the action level. Together, these procedures enabled a systematic evaluation of the LLM pipeline across multiple axes: input representation, inference-time context, temporal robustness, and predictive calibration.

#### Evaluation on user-initiated actions

Furthermore, to focus on deliberate clinician behavior rather than actions autogenerated from EHR interface elements, we repeated the analysis using only actions more likely to be user-initiated, defined as actions occurring greater than one second after a previous action event. This approach helped in excluding potentially automated or platform-triggered (i.e., by the Epic EHR) events and provided an additional assessment of intentional action sequences.

## Results

During the study period, there were 2,478,503 orders, and an associated 295,912,142 audit log actions. The cohort used for model training included 1,006,261 action sequences preceding orders placed between 2019.01–2020.07. Specifically, 81,632,666 audit log actions from 711,030 sequences associated with orders (∼70%) were used for model training, 10,469,849 audit log actions from 94,203 sequences associated with orders (∼10%) used to determine early stopping criteria, and 24,052,295 audit log actions from 201,028 sequences associated with orders (∼20%) were held out for evaluation (*early-period test set*). This period included 1,398 unique ordering clinicians, with 301 (22%) clinicians overlapping across the training and held-out testing sets.

For the two additional temporal validation sets, activity sequences from temporally distinct 1-year periods were used (OOS-1 [2021.06–2022.05]: 673,981 sequences corresponding to orders and 77,628,214 audit log actions; OOS-2 [2023.06–2024.05]: 798,276 sequences corresponding to orders and 102,129,118 audit log actions). Across each period, there were similar proportions of overlapping clinicians between the training and OSS datasets (OOS-1: 338 [24%], OOS-2: 224 [16%]).

The demographic and descriptive characteristics for the cohorts and audit log action sequences included in this study are shown in Table 1.

**Table 1.** Description of datasets used for model training and evaluation. The early-period test set was drawn from the same temporal window as the training data (2019.01–2020.07). Robustness to temporal drift was evaluated using two action sequence datasets drawn from temporal hold-out time periods: OOS-1 (2021.06–2022.05), and OOS-2 (2023.06–2024.05).

| Dataset | Train | Test | OOS-1 | OOS-2 |
| --- | --- | --- | --- | --- |
| Period | 2019.01 – 2020.07 |  | 2021.06 – 2022.05 | 2023.06 – 2024.05 |
| # Included Clinicians |  |  |  |  |
| *Physicians* | 797 (69.9%) | 320 (67.4%) | 594 (57.0%) | 442 (39.5%) |
| *APPs* | 282 (24.7%) | 128 (26.9%) | 280 (26.8%) | 310 (27.7%) |
| *Residents & Fellows* | 31 (2.7%) | 17 (3.6%) | 137 (13.1%) | 328 (29.3%) |
| *Others (Unknown)* | 31 (2.7%) | 11 (2.1%) | 32 (3.1%) | 38 (3.4%) |
| # EHR Action Sequences Associated with Orders | 711,030 | 201,028 | 673,981 | 798,276 |
| Action Sequence Length |  |  |  |  |
| *25th* | 51 | 52 | 58 | 63 |
| *50th* | 87 | 89 | 96 | 105 |
| *75th* | 140 | 143 | 148 | 165 |
| # Total Actions | 81,632,666 | 24,052,295 | 77,628,214 | 102,129,118 |
| # Inferred Actions | N/A | 20,461,603 | 69,651,774 | 92,871,386 |
| # Inferred Actions (User-Initiated) | N/A | 11,289,131 | 46,317,666 | 54,516,811 |

### Evaluation on early-time period

For the early-period test set, the word-based LLM achieved the best accuracy at 0.7418 (95% CI: 0.7415–0.7420) compared to both the field-based LLM (accuracy: 0.3838, 95% CI: 0.3836–0.3840) and the Markov baseline (accuracy: 0.4553, 95% CI: 0.4551–0.4555), as shown in Table 2.

**Table 2.** Model performance on the early-period test sample. This in-sample set consisted of a 20% sampling from the same period as the training set (2019.01–2020.07), randomly selected and held-out entirely during the fine-tuning process. Prediction of the next EHR action was performed over a total of 20,461,603 EHR actions. 95% confidence intervals (CI) were generated from 1000 bootstrap resampling iterations. Accuracy is shown for predicting the observed next action, and also for whether the observed next action was within the top 5 model output choices (Accuracy @ Top-5).

| Experiment | Accuracy (95% CI) | Accuracy @ Top-5 (95% CI) |
| --- | --- | --- |
| **Markov Baseline** | 0.4553 (0.4551–0.4555) | 0.7971 (0.7969–0.7972) |
| **Field-based LLM** | 0.3838 (0.3836–0.3840) | 0.8233 (0.8232–0.8235) |
| **Word-based LLM<sup>a</sup>** | 0.7418<sup>b</sup> (0.7415–0.7420) | 0.8575 (0.8573–0.8577) |

<sup>a</sup> For the word-based LLM, prediction of the next EHR action was performed over a total of 13,397,377 (65.4757%) actions due to truncation to fit LLM’s context window limits.
<sup>b</sup> Accuracy after post-hoc nearest neighbor action label matching. Raw accuracy before matching was 3.4029e-4 (3.3111e-4–3.4007e-4).

### Evaluation on temporal hold-out samples

Across both out-of-sample evaluation periods, the word-based LLM consistently outperformed the Markov baseline (Table 3). In OOS-1, the word-based LLM achieved an accuracy of 0.6509 (95% CI: 0.6508–0.6511) compared to the Markov baseline (accuracy: 0.3169, 95% CI: 0.3168–0.3170). A similar trend was observed in OOS-2, with the word-based LLM showing an accuracy of 0.6232 (95% CI: 0.6231–0.6233) compared to the Markov baseline (accuracy: 0.2648, 95% CI: 0.2647–0.2649).

**Table 3.** Model performance on the temporal hold-out validation sets. To assess model’s generalizability to action sequences from temporally distinct action sequences, we also evaluated the model on two out-of-sample test sets drawn from future time periods: OOS-1 (2021.06–2022.05; 69,651,774 EHR actions), and OOS-2 (2023.06–2024.05; 92,871,386 EHR actions). The best-performing LLM model (i.e., Word-based LLM) was compared against the non-contextual Markov baseline approach. 95% confidence intervals (CI) from 1000 bootstrap resampling iterations is reported across all metrics.

| Test Period | Model Configuration | Accuracy (95% CI) | Accuracy @ Top-5 (95% CI) |
| --- | --- | --- | --- |
| **OOS-1** | Markov Baseline | 0.3169 (0.3168–0.3170) | 0.6356 (0.6355–0.6358) |
|  | Word-based LLM<sup>a</sup> | 0.6509<sup>b</sup> (0.6508–0.6511) | 0.8030 (0.8029–0.8031) |
| **OOS-2** | Markov Baseline | 0.2648 (0.2647–0.2649) | 0.5166 (0.5165–0.5167) |
|  | Word-based LLM<sup>c</sup> | 0.6232<sup>d</sup> (0.6231–0.6233) | 0.7660 (0.7659–0.7660) |

<sup>a</sup> For the word-based LLM in OOS-1, prediction of the next EHR action was performed over a total of 45,630,757 (65.5127%) actions due to truncation to fit LLM’s context window limits.
<sup>b</sup> Accuracy after post-hoc nearest neighbor action label matching. Raw accuracy before matching was 3.0670e-4 (3.0155e-4–3.1117e-4).
<sup>c</sup> For the word-based LLM in OOS-2, prediction of the next EHR action was performed over a total of 55,287,088 (59.5308%) actions due to truncation to fit LLM’s context window limits.
<sup>d</sup> Accuracy after post-hoc nearest neighbor action label matching. Raw accuracy before matching was 2.4738e-4 (2.4317e-4–2.5174e-4).

Additional metrics regarding model performance are provided in Supplementary Material (Section A3).

### Evaluation on user-initiated actions

The word-based LLM also outperformed the Markov baseline when sequences were filtered to only include actions more likely to be user-initiated (Table 4). In the early-period test set, the LLM achieved an accuracy of 0.6596 (95% CI: 0.6594–0.6599) compared to the Markov baseline (0.3416, 95% CI: 0.3413–0.3419). Similar performance was observed in both OOS-1 and OOS-2, with the LLM showing an accuracy of (OOS-1: 0.6005 [0.6003–0.6006], OOS-2: 0.5723 [0.5722–0.5725]) compared to the Markov baseline (OOS-1, 0.0430 [0.0429–0.0431]; OOS-2, 0.0843 [0.0843–0.0844]).

**Table 4.** Comparison of model performances on inferring user-initiated action subset, across all evaluation sets. Action sequences was restricted to consecutive actions that occurred at least 1 second apart, as these events were considered to be more likely to be initiated by the user (vs autogenerated actions from the EHR; early-period set: 11,289,131 actions, OOS-1: 46,317,666 actions, OOS-2: 54,516,811 actions). 95% confidence intervals (CI) from 1000 bootstrap resampling iterations are reported across all metrics.

| Test Period | Model Configuration | Accuracy (95% CI) | Accuracy @ Top-5 (95% CI) |
| --- | --- | --- | --- |
| **Early-Period Test Set**<br>(2019.01–2020.07) | Markov Baseline | 0.3416 (0.3413–0.3419) | 0.7252 (0.7250–0.7255) |
|  | Word-based LLM<sup>a</sup> | 0.6596<sup>b</sup> (0.6594–0.6599) | 0.8045 (0.8043–0.8048) |
| **OOS-1**<br>(2021.06–2022.05) | Markov Baseline | 0.0430 (0.0429–0.0431) | 0.1916 (0.1915–0.1917) |
|  | Word-based LLM<sup>c</sup> | 0.6005<sup>d</sup> (0.6003–0.6006) | 0.7644 (0.7643–0.7645) |
| **OOS-2**<br>(2023.06–2024.05) | Markov Baseline | 0.0843 (0.0843–0.0844) | 0.1677 (0.1676–0.1678) |
|  | Word-based LLM<sup>e</sup> | 0.5723<sup>f</sup> (0.5722–0.5725) | 0.7374 (0.7373–0.7375) |

<sup>a</sup> For the word-based LLM in the early-period test set, prediction of the next EHR action was performed over a total of 11,128,162 (98.5741%) actions due to truncation to fit LLM context window limits.
<sup>b</sup> Accuracy after post-hoc nearest neighbor action label matching. Raw accuracy before matching was 1.1941e-3 (1.1727e-3–1.2139e-3).
<sup>c</sup> For the word-based LLM in OOS-1, prediction of the next EHR action was performed over a total of 40,916,505 (88.3389%) actions due to truncation to fit LLM context window limits.
<sup>d</sup> Accuracy after post-hoc nearest neighbor action label matching. Raw accuracy before matching was 8.9710e-4 (8.8793e-4–9.0499e-4).
<sup>e</sup> For the word-based LLM in OOS-2, prediction of the next EHR action was performed over a total of 47,623,342 (87.3553%) actions due to truncation to fit LLM context window limits.
<sup>f</sup> Accuracy after post-hoc nearest neighbor action label matching. Raw accuracy before matching was 8.0658e-4 (7.9831e-4–8.1450e-4).

## Discussion

Using a large data set that included >295 million clinician actions associated with >2.4 million clinician orders, we systematically evaluated how an LLM could be designed to model the trajectories of clinician-EHR interactions. Our primary finding was that a semantic, word-based LLM substantially outperformed both a symbolic, field-based LLM, and a first-order Markov baseline for next-action prediction. This performance advantage persisted across two temporally distinct out-of-sample periods, suggesting that the semantic, word-based LLM was able to learn the recurring structure in audit log action sequences, consistent with clinician-EHR interaction patterns, that generalized beyond the original training period.

Our central methodological finding was that the semantic representation of audit log action sequences performed substantially better than the symbolic representation, suggesting that the human readable descriptions attached to audit log actions contained useful information for modeling clinician workflow. Prior applications of NLP techniques to audit log sequences have treated actions as discrete tokens, similar to the symbolic LLM approach.^35–37^ Our results suggest that preserving a semantic layer in action descriptions can improve the ability of sequence models to learn reusable action sequences. This is notable because vendor-generated action labels – as is the case with audit logs – are often noisy, inconsistent, and difficult to interpret without clinical and EHR expertise. Despite these limitations, the semantic approach appeared to be able to use information in the text labels to improve prediction and temporal generalization.

The semantic model’s advantage was also observed when evaluation was restricted to actions that were likely to be user-initiated, suggesting that the observed performance gains were not solely attributable to memorization of repetitive system-generated events. At the same time, free-text generation using the semantic approach rarely produced the exact valid audit log action label. In our analytical framework, the semantic LLM’s raw output required mapping to the nearest valid action description, suggesting that semantic models may be most useful when paired with a constrained action vocabulary and retrieval-based decoding to ensure outputs correspond to valid EHR actions. Taken together, these findings suggest that the choice of EHR action representation (i.e., semantic vs symbolic) may be at least as important as the sequence-modeling architecture itself (i.e., Llama3) when applying language models to EHR audit logs. Another notable feature of the semantic model was its stronger performance in temporally distinct evaluation periods.

Audit log data are often subject to temporal drift because of changes to EHR interfaces, shifts in clinician roles and staffing patterns, redesigned workflows, and evolving clinical practice. The observed decline in performance across OOS periods indicated that temporal drift remained a potential challenge. However, the semantic model degraded less than the Markov baseline, suggesting that semantic representations may provide robustness when exact action transitions change over time. This is relevant because models trained in a period ideally would remain informative after EHR upgrades, order-set modifications, staffing changes, or workflow redesigns. Semantic model performance remained relatively robust despite substantial differences between the training and out-of-sample datasets, including shifts in clinician composition over time. These results further suggest that the learned representations generalized across distributional changes (e.g., different clinician groups, expertise) rather than merely memorizing action patterns specific to the training cohort. For example, changes in the relative proportions of physicians, advanced practice providers, residents, and fellows over time could contribute to differences in action sequence distributions and shifts in model performance.

Therefore, some of the observed temporal degradation may reflect shifts in user mix rather than EHR interface or workflow changes alone. Future work should evaluate performance stratified by clinician role, specialty, care setting, and order type, and should examine whether role-specific or specialty-specific sequence models can capture representations aligned to their clinical work activities.

The relatively poor performance of the symbolic LLM compared with the Markov baseline warrants further consideration. One plausible explanation is that the symbolic action representation did not leverage the pretrained model’s existing linguistic knowledge. Because the symbolic approach used action identifier tokens (e.g., METRIC IDs from the audit log actions), it required the symbolic LLM to learn relationships among actions from the audit log data alone rather than benefiting from semantic information acquired during large-scale language pretraining. Under these conditions, the advantages of a pretrained transformer architecture may have diminished. Therefore, sparse or infrequent action event transitions may have been easier for a Markov model to memorize (as local bigram probabilities) than for the symbolic LLM to learn through fine-tuning. Moreover, the hyperparameters and tokenizer design may have been better suited to natural language tokenization than to a purely symbolic action vocabulary. These findings suggest that symbolic modeling of audit log actions may require architectures or pretraining strategies specifically designed for discrete event sequences.

Although we evaluated model performance on the next-action prediction task, our intention was not to suggest that it is useful to predict every clinician action in real time. Rather, next-action prediction served as an objective benchmark for evaluating whether a model had learned meaningful action sequence structure from raw audit log sequences. A model that anticipates the next EHR action from the available prior context has likely encoded reasonable representations of recurring clinical tasks, common navigation paths, and expected transitions between EHR actions. These learned representations can support future applications that require understanding the structure of clinical work, such as identifying task segments from unlabeled audit logs, monitoring workflow changes after EHR interface modifications, comparing ordering workflows across clinician roles or settings, identifying inefficient navigation patterns, or studying action sequences that precede order modification, cancellation, wrong-patient risk, or other safety-relevant events.

This work was inspired by research in human-computer interaction (HCI) that characterizes user interaction patterns as being structurally similar to human language,^8,53^ with latent structural patterns – or a *grammar* – reflecting user strategies and preferences in performing routine tasks.^54,55^ However, much of the past work in HCI, or in studies using audit logs^35–37^ have not made meaningful attempts to determine whether human action sequences have inherent underlying sequential structure, similar to a language. As such, the framework and developed analytical approach has several implications for clinical informatics research. First, it suggests that audit logs can be studied not only as sources of aggregate workload measures but also as sequential records of clinical work. Second, it demonstrates that preserving natural language action descriptions may improve sequence modeling compared with treating actions as arbitrary symbolic identifiers. Third, it provides a scalable framework for evaluating workflow representations using temporally separated validation periods, which is important because EHR-based workflows are not static. Finally, as previously described, this framework offers a path toward transforming low-level audit log event streams into reusable workflow representations that can support operational monitoring, EHR design evaluation, and safety-focused informatics research.

Finally, our algorithmic framework is modular and flexible, meaning it can be extended in various ways for future improvements. As novel open-weight pre-trained LLM architectures are released (e.g., Llama4, GPT-OSS, Gemma 3, DeepSeek V3.2), they can easily be swapped into our pipeline for improved predictive performance. The LLM pipeline could also be extended to learn from metadata about the action using a tabular approach. For example, by including the clinician ID or patient identifier associated with each action, the LLM could be designed to learn individual-level interaction behaviors or anticipate consecutive actions that belong to the same patient’s care (e.g., a role-specific or a specialty-specific model). Future work could also examine how the symbolic and semantic LLM approaches could be integrated within a single model. Apart from our work focusing on EHR interaction sequences, a similar LLM pipeline could be designed to learn from longitudinal sequence of clinical data documented for each encounter; for example, a tabular sequence of EHR documentation activities along with the exact value recorded (e.g., diagnosis code, vitals measurement, prescription, or clinical note content) could be tokenized and trained using an LLM for various classification tasks (e.g., set of most relevant codes that should be billed for an encounter).^52^

Several limitations should be noted. First, this study was conducted at a single academic health system using inpatient audit logs, and the findings may not generalize to other institutions, EHR configurations, specialties, or ambulatory settings. Second, the model was trained only on action sequences preceding orders. This design was focused on a common and clinically consequential workflow, but it does not capture all EHR-mediated clinical activities. Third, the word-based LLM was evaluated on substantially fewer actions than the symbolic and Markov approaches (e.g., 13,397,377 vs 20,461,603 actions, ∼65%, in the early-period test set). Because each action description fragments into multiple subword tokens, far fewer actions fit within the fixed context window, and because sequences were truncated to their most recent tokens, the earliest actions of long sequences were dropped from the input and were not scored. For the symbolic and Markov models, where each action corresponds to approximately one token, nearly all actions fit within the context window and were evaluated. Because the excluded actions for the semantic model were systematically the earliest actions of the longest sequences, the semantic model was scored preferentially on shorter sequences and on the later portions of longer sequences; if action distributions differ by position within a sequence, this could bias the cross-model comparison.

However, this asymmetry does not favor the word-based model: on the actions it was scored on, it operated with substantially less action-level context than the field-based model, because each action description consumed multiple subword tokens, yet it outperformed both baselines by a large and consistent margin across all evaluation sets. Fourth, the semantic model depended on vendor-provided action descriptions, which may be inconsistent, ambiguous, or institution-specific. Finally, LLM-based modeling required substantially greater computational resources than the simpler baselines. Finally, there were changes in the clinician composition in the OOS periods, which may have impacted the model performance changes in these periods.

### Conclusions

In conclusion, semantic sequence modeling of EHR audit logs can learn recurring structure in clinician-EHR workflows and can outperform symbolic action modeling and minimal-context transition models for next-action prediction. These findings suggest that human-readable audit log action descriptions contain useful information for modeling clinical work and that longer-context sequence models can capture workflow structure from high-volume EHR event streams. By reframing audit logs as ordered representations of clinical activity – or actions-as-language – rather than only as sources of aggregate workload measures, this approach provides a foundation for future informatics applications in task identification, workflow optimization, monitoring of EHR changes, clinician-burden measurement, and safety-focused analyses of potentially error-prone workflow transitions.

## Author contributions

SK, SSL and TK conceptualized the idea; SK, SSL, AC, SJ and TK were involved in the design of the experiments; SK, TK and SSL wrote the initial version of the manuscript. All authors substantially edited the manuscript and provided approval for the final version.

## Data Availability

All data produced in the present study are available upon reasonable request to the authors

