## Supplement for "A language model framework for sequence modeling of EHR audit logs to characterize clinician-EHR interactions"

**Table A1**. Model card for the models discussed.

| **Model Details** | |
| --- | --- |
| Model Developers | Washington University in St. Louis |
| Input | Epic EHR audit log data, up to 512 tokens |
| Output | Tokens representing generated Epic EHR audit log events or its subwords in the METRIC_DESCRIPTION column |
| Model Architecture | Publicly available Llama-3 (8B) by Meta |
| Model Dates | These models were fine-tuned in May and December 2025. |
| Status | This model was fine-tuned with a private dataset of inpatient clinician audit logs. We do not anticipate releasing updates to our model. |
| **Intended Use** | |
| Intended Use Cases | Our fine-tuned models are intended for research use with Epic EHR audit logs for generative modeling. |
| Out-of-Scope Usage | Because of the experimental nature of these models, we do not recommend the use of our models in a non-research context. |
| **Hardware and Software** | |
| Training | We used Hugging Face transformers and Transformer Reinforcement Learning for the model architectures on top of PyTorch,^56^ and we use AdamW^57^ to optimize. We trained on NVIDIA 80GB A100s in a research compute cluster maintained at Washington University in St. Louis. |
| **Training Data** | |
| Overview | These models were trained on a set of audit logs obtained from selected inpatient clinicians performing work in the Barnes-Jewish Hospital and Washington University School of Medicine as part of the IGNITE study (IRB#202209008; Agency for Healthcare Research and Quality (AHRQ) (Grant #R01HS029020)). The original training data is protected by health information (PHI) and cannot be released. |
| Data Freshness | The training data was taken over a period of January 2019 to July 2020. |
| **Evaluation Results** | |
| See the Results Section for details on the performance of each of our models. | |
| **Ethical Considerations and Limitations** | |
| Because of their experimental nature, we do not recommend the usage of these models outside of a research context. In addition, no generated data will contain PHI; however, it contains proprietary event descriptors from Epic EHR system. | |

### A1. LLM Fine-Tuning

In the field-based approach, we identified the 100 most frequent actions across the training set and assigned each a unique symbolic token (e.g., Action 1, Action 2). These tokens were added to the tokenizer’s vocabulary, while all remaining less frequent actions were grouped into a single rare-action token. This threshold provided a practical balance that captures the majority of routine behavioral transitions while avoiding excessive sparsity and overfitting to highly infrequent or variable actions.

Fine-tuning on domain-specific data enabled the model to adapt to the unique patterns of clinician EHR interaction while preserving the pretrained model’s capacity to learn temporal dependencies, sequence progression, and general language-based reasoning. Starting with a pretrained backbone substantially reduces the burden of learning sequential dynamics from scratch and supports more efficient and stable adoption to the EHR activity domain.

Fine-tuning all parameters of an LLM is often unnecessary and computationally expensive, especially when adapting to a domain-specific task with modest training corpus. To address this, we incorporated Parameter-Efficient Fine-Tuning (PEFT) methods. Specifically, we used Low-Rank Adaptation (LoRA),^13^ which introduces small trainable low-rank matrices into otherwise frozen pretrained weights. During fine-tuning, only these adapter layers are updated, enabling the model to learn domain-specific behavior while preserving its general sequence-modeling capabilities. This substantially reduces memory usage and training time and supports a modular architecture in which separate LoRA adaptors can be trained for different clinical tasks without retraining the entire model.

To further reduce the GPU memory requirements, we applied model quantization during fine-tuning. Quantization lowers the precision of model weights, allowing large models to be trained and deployed on resource-constrained hardware with minimal performance degradation. We adopted 4-bit quantization in combination with LoRA (QLoRA), using default configurations recommended in the original implementation.^13^ This approach enabled us to fine-tune Llama-3-8B efficiently on commodity hardware while maintaining stable training dynamics.

During inference on the held-out datasets, we incorporated a short warm-up context prompt to improve prediction stability and contextual alignment. This prompt provided the model with a small number of correct initial tokens to anchor its sequential reasoning at the start of each evaluation sequence.
Because prompting operates entirely at the input level, it enhances the model’s behavior at inference time without modifying the underlying weights.

### A2. Evaluation Comparisons

**Table A2.** Summary of evaluation comparisons for the next-action prediction task. All conditions were evaluated over identical token positions in the test sets. All LLM hyperparameter and model configurations were kept the same between the field-based and word-based approaches.

| **Condition** | **Input Format** | **Description** |
| --- | --- | --- |
| Markov Baseline | Single previous action | First-order Markov model using bigram transition probabilities observed from training data |
| Field-based LLM | 30-minute history of symbolic tokens | LLM fine-tuned on EHR actions mapped to symbolic action categories (e.g., [action 1], [action 2], ...) |
| Word-based LLM | 30-minute history of semantic descriptions | LLM fine-tuned on natural language words from the text labels given to EHR actions (e.g., “Order placed”, “Chart viewed”, ...) |
| User-Initiated Action Modeling | 30-minute history of actions tokenized in either way | LLM fine-tuned on EHR action sequences that included only actions occurring more than one second after the previous action |

### A3. Model Evaluation

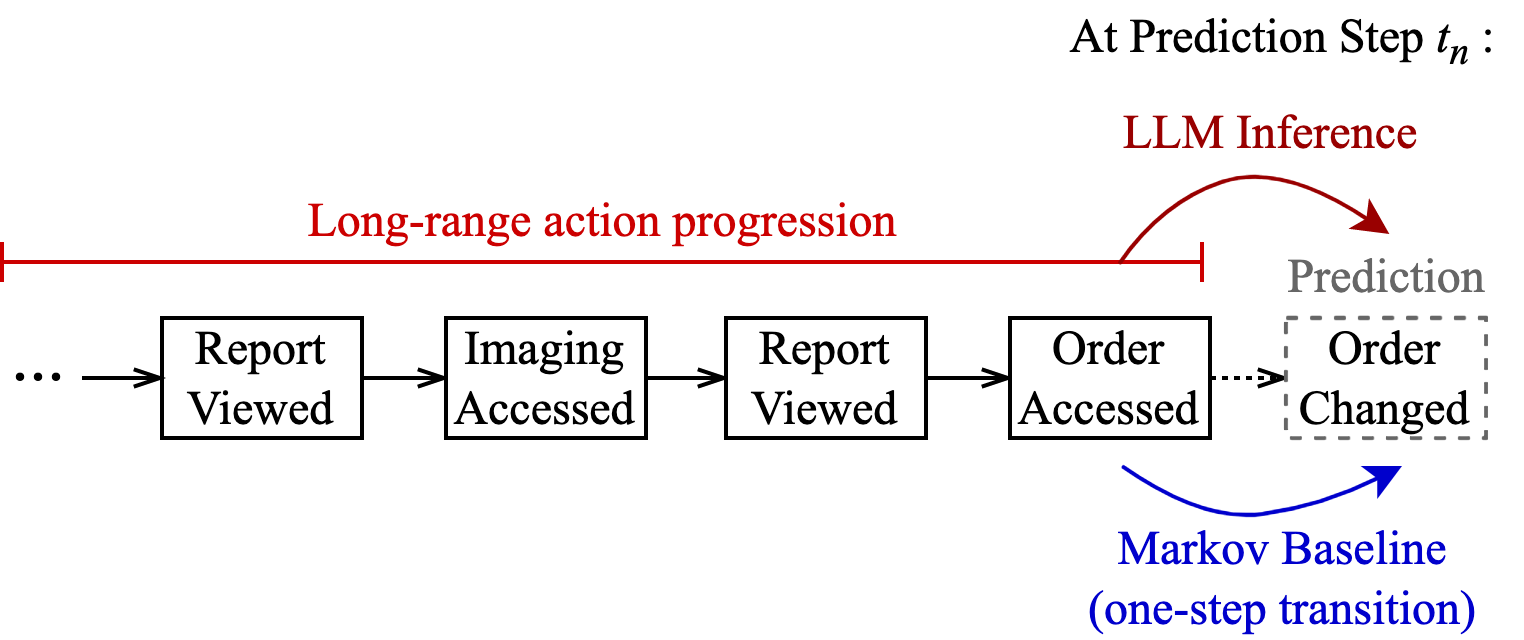

**Figure A1.** Illustration of sequential learning evaluation comparison. LLM-based method leverages long-range dependencies to predict the next action, whereas a first-order Markov baseline uses simple one-step transition probabilities observed from training data.

At inference time, for the LLM-based models, the first ten correct tokens of each sequence were provided as an initial conditioning window (i.e., warm-up context prompt). To ensure that all models were evaluated over identical positions regardless of whether the warm-up prompt was applied, we excluded the first ten tokens from performance computations in all models.

Action-level accuracy measured the proportion of correctly predicted actions across sequences and directly reflected the model's ability to capture sequential patterns in clinician behavior. Meanwhile, accuracy@top-5 assessed whether the true action appeared among the five highest-ranked candidates. Because word-based LLMs operate at the subword level, we evaluated them at the action level using an aggregation: nearest-neighbor accuracy, which mapped the generated text to the closest real action description. Nearest-neighbor matching enabled a more interpretable ranking and was used for all top-k metrics (i.e., top-1, top-5). In our setup, we used term-frequency inverse-document frequency (TF-IDF) n-gram embeddings (3–5-grams) of the entire set of real action descriptions. We then used the same fitted vectorizer to transform predicted raw action descriptions and computed *l2* cosine similarity scores between each prediction’s embedding and real descriptions’ embeddings to rank nearest neighbor.

Perplexity score measured how efficiently the model distributed probability mass over the correct next actions, which measured the average uncertainty the model has when predicting the true next token in a sequence—lower perplexity penalizes confident but incorrect guesses. Lower perplexity indicated higher confidence and better fit to the observed sequences. For example, a perplexity of 10 can be interpreted as the model narrowing its next prediction to about 10 plausible actions, with uncertainty remaining within that narrowed scope. These metric complements accuracy-based metrics by quantifying the overall likelihood assigned to observed sequences, regardless of whether the top prediction was correct. For word-based LLMs, per-token cross-entropy values were aggregated (mean pooled) to the action level prior to computing perplexity. Specifically, it was computed over subword tokens due to the nature of the different tokenization approach compared to the Markov baseline and field-based LLM’s full action-level tokenization, making its absolute magnitude not directly comparable across models.

To quantify the model’s calibration and uncertainty, we incorporated entropy and expected calibration error (ECE). Entropy quantified the uncertainty of the model’s predicted probability distribution at each step (i.e., *how diffused* the model’s belief is). Higher entropy values indicated less confident or more ambiguous predictions. A mean average of action-level entropies across all predicted actions were computed and reported. ECE quantified model calibration by measuring the alignment between predicted confidence and empirical correctness. This produced a single summary value where lower scores indicate a better-calibrated model, whose probability estimates correspond closely to true correctness rates. ECE was calculated by partitioning predicted probabilities into fixed intervals (e.g. 10 confidence bins) and measuring the weighted difference between predicted confidence and empirical accuracy in each bin. As with perplexity, token-level entropy and confidence estimates for word-based models LLMs were averaged across subword tokens to produce action-level values.

### A3. Additional Model Evaluation Results

The word-based LLM yielded a higher perplexity of 5.4274e-5 (95% CI: 5.4196e5–5.4351e5) and ECE of 0.8054 (95% CI: 0.8054–0.8055) than both alternatives (Table A2).

**Table A2.** Model performance on the early-period test sample. This in-sample set consisted of a 20% sampling from the same period as the training set (2019.01–2020.07), randomly selected and held-out entirely during the fine-tuning process. Prediction of the next EHR action was performed over a total of 20,461,603 EHR actions. 95% confidence intervals (CI) were generated from 1000 bootstrap resampling iterations.

| **Experiment** | **Perplexity Score (95% CI)** | **Mean Entropy (95% CI)** | **ECE Score (95% CI)** |
| --- | --- | --- | --- |
| **Markov Baseline** | 7.1078^a^ (7.1032–7.1130) | 1.9589 (1.9586–1.9592) | 0.0080 (0.0078–0.0082) |
| **Field-based LLM** | 8.0417 (8.0366–8.0475) | 1.7185 (1.7182–1.7189) | 0.1364 (0.1362–0.1366) |
| **Word-based LLM^b^** | 5.4274e5 (5.4196e5–5.4351e5) | 0.7060 (0.7057–0.7062) | 0.8054 (0.8054–0.8055) |

^a^ 628 (0.0031%) token positions with unseen transitions were skipped from computing the perplexity score (no probability value exists in the 1^st^-order Markov transition matrix)

Across both out-of-sample evaluation periods, the word-based LLM consistently outperformed the Markov baseline. In OOS-1, the LLM achieved a lower entropy of 0.8397 (95% CI: 0.8395–0.8398) compared to the Markov baseline (Table A3). A similar trend was observed in OOS-2, with the LLM showing a lower entropy of 1.0022 (95% CI: 1.0020–1.0023) compared to the Markov baseline. However, as in the early-period evaluation, the LLM showed a higher perplexity (OOS-1: 3.4136e5 [95% CI: 3.4112e5–3.4160e5], OOS-2: 1.9185e5 [95% CI: 1.9172e5–1.9198e5]) and ECE (OOS-1: 0.7716 [95% CI: 0.7715–0.7716], OOS-2: 0.7421 [95% CI: 0.7420–0.7421]) compared to the Markov baseline in both periods.

**Table A3.** Model performance on the temporal hold-out validation sets. To assess model’s generalizability to action sequences from temporally distinct action sequences, we also evaluated the model on two out-of-sample test sets drawn from future time periods: OOS-1 (2021.06–2022.05; 69,651,774 EHR actions), and OOS-2 (2023.06–2024.05; 92,871,386 EHR actions). The best-performing LLM model (i.e., Word-based LLM) was compared against the non-contextual Markov baseline approach. 95% confidence intervals (CI) from 1000 bootstrap resampling iterations.

| **Test Period** | **Model Configuration** | **Perplexity Score (95% CI)** | **Mean Entropy (95% CI)** | **ECE Score (95% CI)** |
| --- | --- | --- | --- | --- |
| **OOS-1** | Markov Baseline | 12.4173^a^ (12.4113–12.4231) | 2.2567 (2.2565–2.2568) | 0.0713 (0.0712–0.0714) |
|  | Word-based LLM^b^ | 3.4136e5 (3.4112e5–3.4160e5) | 0.8397 (0.8395–0.8398) | 0.7716 (0.7715–0.7716) |
| **OOS-2** | Markov Baseline | 16.2035^c^ (16.1942–16.2118) | 2.4726 (2.4724–2.4728) | 0.0843 (0.0843–0.0844) |
|  | Word-based LLM^d^ | 1.9185e5 (1.9172e5–1.9198e5) | 1.0022 (1.0020–1.0023) | 0.7421 (0.7420–0.7421) |

^a^ 6,123,461 (9%) token positions with unseen transitions were skipped from computing the perplexity score (no probability value exists in the 1^st^-order Markov transition matrix)

^b^ For the word-based LLM in OOS-1, prediction of the next EHR action was performed over a total of 45,630,757 (65.5127%) actions due to truncation to fit LLM’s context window limits.

^c^ 20,206,973 (22%) token positions with unseen transitions were skipped from computing the perplexity score (no probability value exists in the 1^st^-order Markov transition matrix)

^d^ For the word-based LLM in OOS-2, prediction of the next EHR action was performed over a total of 55,287,088 (59.5308%) actions due to truncation to fit LLM’s context window limits.

Across all evaluation periods, the word-based LLM continued to substantially outperform the Markov baseline when sequences were filtered to only include actions more likely to be user-initiated. In the early-period test set, the LLM achieved a lower entropy of 0.8579 (95% CI: 0.8576–0.8581) compared to the Markov baseline (Table A4). Similar performance was observed in both OOS-1 and OOS-2, with the LLM showing a lower entropy (OOS-1: 0.9436 [0.9434–0.9437], OOS-2: 1.0915 [1.0914–1.0917]) compared to the Markov baseline.

However, in the early-period test set, the LLM showed a higher perplexity of 1.9196e5 (95% CI: 1.9169e5–1.9222e5) and ECE of 0.8054 (95% CI: 0.8054–0.8055) compared to the Markov baseline. Consistently, across both OOS-1 and OOS-2, the LLM showed higher perplexity and ECE compared to the Markov baseline.

**Table A4.** Comparison of model performances on inferring user-initiated action subset, across all evaluation sets. Action sequences was restricted to consecutive actions that occurred at least 1 second apart, as these events were considered to be more likely to be initiated by the user (vs autogenerated actions from the EHR; early-period set: 11,289,131 actions, OOS-1: 46,317,666 actions, OOS-2: 54,516,811 actions). 95% confidence intervals (CI) from 1000 bootstrap resampling iterations is reported across all metrics.

| **Test Period** | **Model Configuration** | **Perplexity Score (95% CI)** | **Mean Entropy (95% CI)** | **ECE Score (95% CI)** |
| --- | --- | --- | --- | --- |
| **Early-Period Test Set** (2019.01–2020.07) | Markov Baseline | 10.7115^a^ (10.7020–10.7209) | 2.3636 (2.3633–2.3638) | 0.0036 (0.0033–0.0039) |
|  | Word-based LLM^b^ | 1.9196e5 (1.9169e5–1.9222e5) | 0.8579 (0.8576–0.8581) | 0.7521 (0.7520–0.7521) |
| **OOS-1** (2021.06–2022.05) | Markov Baseline | 522.2621^c^ (521.7239–522.7902) | 2.4093 (2.4091–2.4094) | 0.3021 (0.3020–0.3022) |
|  | Word-based LLM^d^ | 1.3214e5 (1.3205e5–1.3224e5) | 0.9436 (0.9434–0.9437) | 0.7336 (0.7336–0.7337) |
| **OOS-2** (2023.06–2024.05) | Markov Baseline | 259.0860^e^ (259.8072–259.3785) | 2.5384 (2.5381–2.5386) | 0.2086 (0.2086–0.2087) |
|  | Word-based LLM^f^ | 8.4345e5 (8.4287e5–8.4400e5) | 1.0915 (1.0914–1.0917) | 0.7126 (0.7125–0.7126) |

^a^ 653 (<1%) token positions with unseen transitions were skipped from computing the perplexity score (no probability value exists in the 1^st^-order Markov transition matrix)

^b^ For the word-based LLM in the early-period test set, prediction of the next EHR action was performed over a total of 11,128,162 (98.5741%) actions due to truncation to fit LLM’s context window limits.

^c^ 10,349,462 (22%) token positions with unseen transitions were skipped from computing the perplexity score (no probability value exists in the 1^st^-order Markov transition matrix)

^d^ For the word-based LLM in the early-period test set, prediction of the next EHR action was performed over a total of 40,916,505 (88.3389%) actions due to truncation to fit LLM’s context window limits.

^e^ 20,217,117 (37%) token positions with unseen transitions were skipped from computing the perplexity score (no probability value exists in the 1^st^-order Markov transition matrix)

^f^ For the word-based LLM in the early-period test set, prediction of the next EHR action was performed over a total of 47,623,342 (87.3553%) actions due to truncation to fit LLM’s context window limits.

Although the LLM help up robust performance in accuracy and predictive entropies, its calibration suffered noticeably. The word-based model, although substantially more accurate than the Markov baseline or field-based LLM, exhibited higher expected calibration error, indicating systematic overconfidence in its predictions. This in part could be due to the LLM demonstrating a consistently lower average entropy than the Markov approach across all scenarios, suggesting that the model formed more confident and concentrated probability distributions around plausible next actions. This trade-off—high predictive performance with poorer calibration—persisted both in early-period and in OSS testing, highlighting calibration as a key challenge for transformer-based behavioral modeling.

Furthermore, while the Markov and field-based LLM show modest perplexity values reflecting action-level predictions, the word-based LLM’s perplexity appears highly inflated because it is computed over subword tokens due to the nature of the different tokenization approach compared to the Markov baseline and field-based LLM’s full action-level tokenization, making its absolute magnitude not directly comparable across models.
